# Regional opening strategies with commuter testing and containment of new SARS-CoV-2 variants in Germany

**DOI:** 10.1101/2021.04.23.21255995

**Authors:** Martin J. Kühn, Daniel Abele, Sebastian Binder, Kathrin Rack, Margrit Klitz, Jan Kleinert, Jonas Gilg, Luca Spataro, Wadim Koslow, Martin Siggel, Michael Meyer-Hermann, Achim Basermann

**Affiliations:** Institute for Software Technology, German Aerospace Center, Cologne, Germany; Department of Systems Immunology and Braunschweig Integrated Centre of Systems Biology (BRICS), Helmholtz Centre for Infection Research, Braunschweig, Germany

**Keywords:** SARS-CoV-2, Covid-19, Nonpharmaceutical intervention, Mitigation strategy, Modeling, Predictive Analytics, NoCovid strategy

## Abstract

**Background:** Despite the vaccination process in Germany, a large share of the population is still susceptible to SARS-CoV-2. In addition, we face the spread of novel variants. Until we overcome the pandemic, reasonable mitigation and opening strategies are crucial to balance public health and economic interests.

**Methods:** We model the spread of SARS-CoV-2 over the German counties by a graph-SIR-type, metapopulation model with particular focus on commuter testing. We account for political interventions by varying contact reduction values in private and public locations such as homes, schools, workplaces, and other. We consider different levels of lockdown strictness, commuter testing strategies, or the delay of intervention implementation. We conduct numerical simulations to assess the effectiveness of the different intervention strategies after one month. The virus dynamics in the regions (German counties) are initialized randomly with incidences between 75-150 weekly new cases per 100,000 inhabitants (red zones) or below (green zones) and consider 25 different initial scenarios of randomly distributed red zones (between 2 and 20 % of all counties). To account for uncertainty, we consider an ensemble set of 500 Monte Carlo runs for each scenario.

**Results:** We find that the strength of the lockdown in regions with out of control virus dynamics is most important to avoid the spread into neighboring regions. With very strict lockdowns in red zones, commuter testing rates of twice a week can substantially contribute to the safety of adjacent regions. In contrast, the negative effect of less strict interventions can be overcome by high commuter testing rates. A further key contributor is the potential delay of the intervention implementation. In order to keep the spread of the virus under control, strict regional lockdowns with minimum delay and commuter testing of at least twice a week are advisable. If less strict interventions are in favor, substantially increased testing rates are needed to avoid overall higher infection dynamics.

**Conclusions:** Our results indicate that local containment of outbreaks and maintenance of low overall incidence is possible even in densely populated and highly connected regions such as Germany or Western Europe. While we demonstrate this on data from Germany, similar patterns of mobility likely exist in many countries and our results are, hence, generalizable to a certain extent.

## Background

With about three million detected infections in Germany [1] and about 22 million vaccinations [2] until April 2021, we could reasonably assume that at least 80% of the population were still susceptible to SARS-CoV-2. New SARS-CoV-2 variants (B.1.1.7, B.1.351 and P.1) contributed to the increase in case numbers [3, Report of Apr. 7]. Due to the higher transmission risk of B.1.1.7 [4], previous mitigation strategies had to be reconsidered and strengthened. Among other things, massive deployment of antigent tests and regular testing were proposed [5].

As of February 2022, we have seen the more infectious Delta and Omikron variants. Although we hoped for herd immunity in winter 2021/2022, still 25 % of the German population have not been vaccinated twice and 43 % of the population have not yet had a third vaccine dose [2]. With lockdowns and strict interventions not being in place, testing becomes even more important.

The prediction of SARS-CoV-2 infections by mathematical models is an active area of research of many groups from all over the world. There are various approaches to simulate the spread of infectious diseases across regions. In the following, we will shortly discuss different approaches and provide a non-exhaustive list of references with applications to SARS-CoV-2.

Sets of ordinary differential equations were already proposed in [6, 7] to model the spread of infectious diseases. These models are often denoted SIR- or SEIR-models and the letters (e.g., *S, E, I*, or *R*) represent infection states used in the model, e.g., *susceptible, exposed, infected*, or *removed/recovered*. In order to avoid a proliferation of letters, we will denote models of this kind as SIR-type models. To be more precise, we denote these models as (deterministic) ODE-SIR-type models to indicate their reliance on ordinary differential equations. A good overview on these deterministic models as well as a presentation of limitations is given, e.g., in [8, 9, 10]. Although having certain limitations, these models are often praised for their simplicity and understandability.

In the context of the SARS-CoV-2 pandemic, ODE-SIR-type models with focus on vaccination were used by [11, 12] while [13] focused on hospitalization and ICU demands. The authors of [14, 15] considered similar models with an additional differentiation of confirmed and unconfirmed infections. This differentiation was also used by the authors of [16, 17]. The authors of [18, 19, 20, 21, 22] considered ODE-SIR models and laid a particular focus on the effect of non-pharmaceutical interventions (NPIs) while [23] focused on contacts outside households and [24] tried to assess the effect of seasonality of SARS-CoV-2. Latency effects in ODE-SIR-type models were studied in [25].

Some obvious limitations of simple ODE-SIR-type models are the homogeneous mixing assumption, the lack of stochastic effects and the implicit use of exponentially distributed compartment stays. The authors of [26] considered stochastic and deterministic ODE-SIR-type models and [27] provides an overview over 13, either stochastic or deterministic, models from 33 papers. The authors of [28] used stochastic compartment models to consider the effect of NPIs and [29] used a stochastic branching process which may be advantageous over compartment models in the beginning phase of pandemic. [30] proposed a simple generalized-growth model for the early phase of disease outbreaks. In [31], different stochastic and deterministic compartment and agent-based models for Germany and Poland from, i.a., [32, 33, 16, 34, 35, 36], were compared.

Stochastic effects such as superspreading events can naturally be modeled by agent-based models or, in parts, by stochastic differential equations. However, the nature and setting of superspreading events is still an area of active research, cf., e.g., [37, 38] and will take years to be fully understood [39]. Agent-based models (ABMs) have been used by many authors since they model infection dynamics in a natural way [40, 41, 42, 43, 44, 45, 46, 47, 48]. Another ABM based on a traffic simulation and mobile phone data was proposed by [49] and [50, 51] presented agent-based models which build upon a predefined contact networks. While ABMs do not have the limitations of the homogeneous mixing assumption or the lack of stochastic events, their use comes at a huge computational overhead.

In [52, 53] agent-based models were combined with artificial intelligence and machine learning. Other machine learning approaches were presented by, e.g., [54, 55, 56]. [54] used ODE-SIR-type based Bayesian inference with invertible neural networks. In [55], an ODE-SIR-type model based machine learning approach was presented and [56] proposed the use of LSTM networks.

Although compartment and agent-based models seem to be contrary approaches, links can be established. The authors of [57, 58] provide links between microscopic, agent-based and ODE models and [59] presented a generalization of ODE-SIR-type models and considered agent-based modeling. In [60], relations between Agent-based and stochastic as well as deterministic metapopulation models were presented.

Metapopulation models reduce the limitations of simple ODE-based models by introducing a spatial dimension and thus allowing for heterogeneous mixing across regions. Metapopulation models were already used before Covid-19 [61] and different approaches exist to extend ODE-models to account for exchange across regions [10, Chapter 14]. The authors of [36, 62, 63, 64, 65] presented ODE-based metapopulation models considering different regions and an additional focus on regional differences in vaccination progress can be found in [66]. [67] presented network-driven contagion phenomena based on ODE-SIR-dynamics but where the latter are not essential for the results provided. Also, agent- and compartment-based models can be combined to set up hybrid models [68].

In order to overcome another limitation of simple ODE-based models, integro-differential equation-based (also named *age of infection*) models [6, 69, 70, 71] can be used. Integro-differential equation-based (IDE) models allow for using arbitrary distributions to waive the implicit use of exponential compartment stays as given by ODE-based models. These have been used for SARS-CoV-2 in, e.g., [72, 73, 74]. A good overview is given in [75, 76]. A trade-off between simple ODE- and IDE-based models are delay-differential equations and linear chain trick [77] also recently used in [78]. The authors of [79] presented memory-equation-based spatial infection dynamics.

The previous background provides a nonexhaustive list of models and papers for mathematical epidemiology. For a broad overview, we refer to [80, 81, 10] and the references therein.

A proactive approach to fight SARS-CoV-2 in Germany and Europe is presented in [82] with the aim of a safe and sustainable re-opening of societies and economies; see also related discussions in [83, 84, 85]. Lockdowns remain implemented regionally until the incidence is below 12 cases per week and 100,000 inhabitants and local measures are reintroduced rapidly should infections flare up again. This approach was already successfully implemented in Australia. It is based on the observation that the virus spreads heterogeneously: There are certain regions with very low incidences while other regions (“hotspots”) are highly affected [3]. However, neighboring regions can quickly become impacted, especially, by daily commuting. So far, the feasibility and effectiveness of the strategy with respect to commuter testing and local lockdowns has not been investigated numerically. The aim of this paper is a quantification of the necessary test frequency, the required strength of the local lockdown and the time frame that we have for the intervention implementation to avoid the spreading of the virus to neighboring regions.

## Methods

The aim of this study is to provide viable strategies of careful opening of facilities in low-incidence regions without being affected by neighboring regions of substantially higher incidence. Motivated by [82], the regions (here: German counties) are partitioned into red and green zones. A region is labeled a *green zone* if there is a stable low incidence below 12 per week and 100,000 inhabitants^[1]^ with effective tracing of new cases. As soon as this is no longer the case, a region is labeled a *red zone*.

We initialize the set of German counties randomly with weekly incidences per 100,000 inhabitants of around 5 for green zones and 75-150 for red zones. The values of 5 and 75-150 are to some extent arbitrary and are chosen such that we have two well distinguished infection dynamics. They are motivated by the proposed strategy [82] and represent infection dynamics which are well under control (green zone) as well as infection dynamics that are out of control and where a lot of infections happen undetected (red zone). We consider 25 different random scenarios with in between 2 and 20 % of the counties as red zones; cf. Fig. 3 (top). Random variables are sampled from a uniform distribution. In green zones, all facilities are open with only protective measures, such as distancing and face masks, in place. Red zones start from an incidence of 12, and in that case a lockdown is implemented for 30 days. For counties with incidence 100 or higher enforced measures are applied. Since political decisions require time, intervention implementation allows for certain delay. Commuter testing is conducted for all commuters coming from red zones. If tested positive, presymptomatic, asymptomatic or symptomatic people will be isolated and prevented from traveling or commuting before recovery. For the precise set of values for lockdowns or testing rates, see the section on *mitigation and opening strategies*. To assess the impact of the initial overall incidence, we also present two scenarios with 42 % and 60 % of counties classified red at the beginning.

Note that throughout this work, incidence always refers to the incidence averaged over 100 000 people and 7 days.

### Mathematical model

In order to achieve the numerical investigation, a large number of ensemble runs of a regionally resolved model have to be conducted. While agent-based models come at a huge computational cost and are well suited for capturing microscopic effects, age-resolved metapopulation models reliably allow for capturing macroscopic effects of a large number of regions at a reasonable cost. We have also decided to use deterministic metapopulation models to avoid making assumptions on the quantification and nature of stochastic events. While this may be a limitation on the county-scale, this will capture the mean macroscopic effects well.

Our mathematical model for the spread of SARS-CoV-2 accounts for age-dependence [86, 87, 88], heterogeneous spread across regions [3], and commuter testing for mitigation [5].

For the geographic resolution, we use a graph approach as presented in [64] and assign one SIR-type model to each German county. The resulting models will be coupled by the edges of the graph which represent the mobility between the regions. The mobility data is obtained from the German Federal Employment Agency [89] complemented with geo-referenced Twitter data [90]. For more details on the data and the practical exchange between regions, we refer to [64]. In the graph, counties are not only connected to their geographical neighbors. We define neighbors by mobility, depending on the numbers of in-commuters. If the number of daily in-commuters in county A (coming from county B) exceeds 1000 on average, B will be classified a neighbor of A. This leads to an increased number of neighbors for larger cities in particular as shown in Fig. 1.

**Figure 1.**
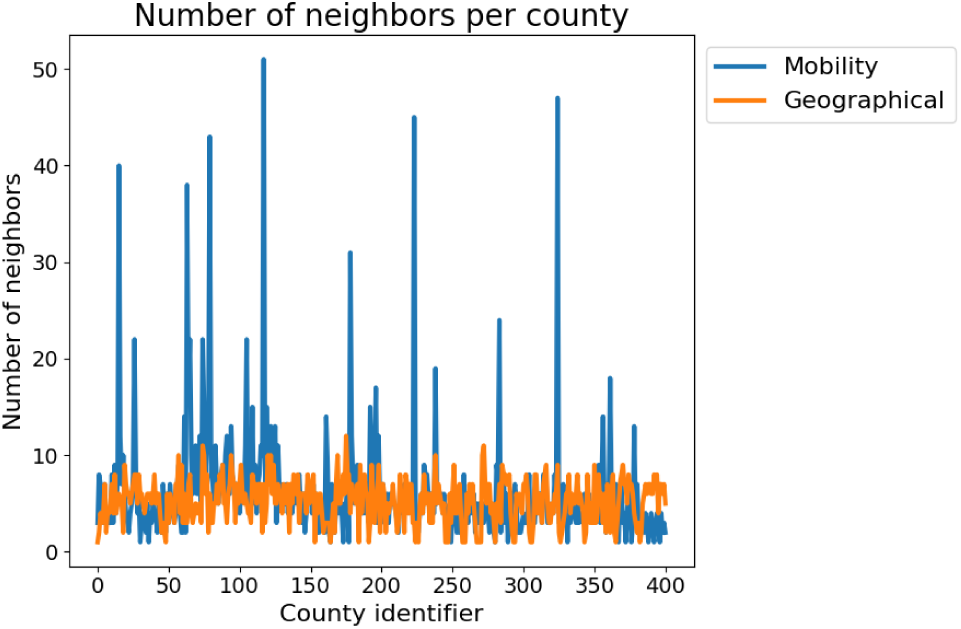
Number of neighboring zones according to geographical and mobility definitions.

For each county, we use an age-resolved SIR-type model based on [14, 64], with particular new focus on commuter testing. The model uses the compartments *Susceptible* (*S*_*i*_); *Exposed* (*E*_*i*_), who carry the virus but are not yet infectious to others; *Carriers* (*C*_*i*_), who carry the virus and are infectious to others but do not yet show symptoms (they may be pre- or asymptomatic); *Infected* (*I*_*i*_), who carry the virus, are infectious and show symptoms; *Hospitalized* (*H*_*i*_), who experience a severe development of the disease; *In Intensive Care Unit* (*U*_*i*_); *Dead* (*D*_*i*_); and *Recovered* (*R*_*i*_), who cannot be infected again. To resolve age-specific disease parameters, the totality of people *N* into *n* different age groups *i* = 1, …, *n*. Note that our focus in this paper is not on hospitalizations, ICU bottlenecks, deaths or vaccinations such that these compartments are of minor interest for our considerations here.

In order to model commuter testing, we introduce the new compartments 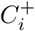 and 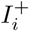 for carriers or infectious who are tested positive while commuting. The compartment 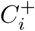 does not have any natural influx and only depends on the number of commuters and testing rates defined between counties on a daily basis. 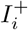 has only influx from 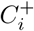 and can also increase due to testing results. For a visualization of the process see Fig. 2. Testing within counties and lockdown strictness will be handled as presented in [64], adapted to counterstrategies against novel SARS-CoV-2 variants. The full system of equations writes

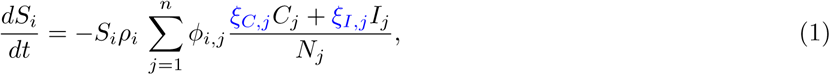

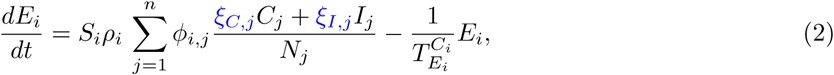

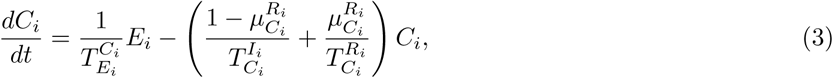

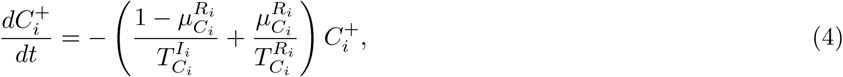

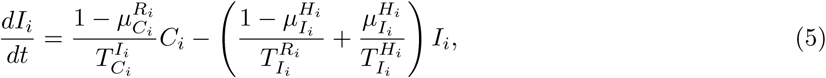

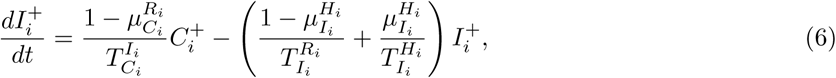

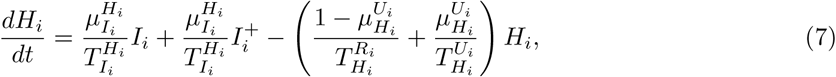

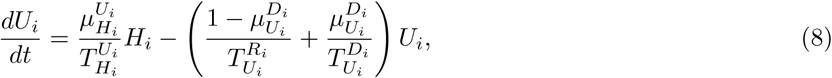

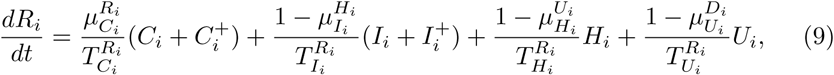

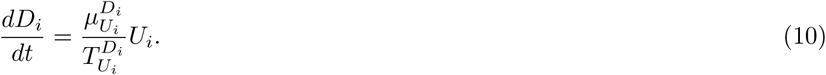

**Figure 2.**
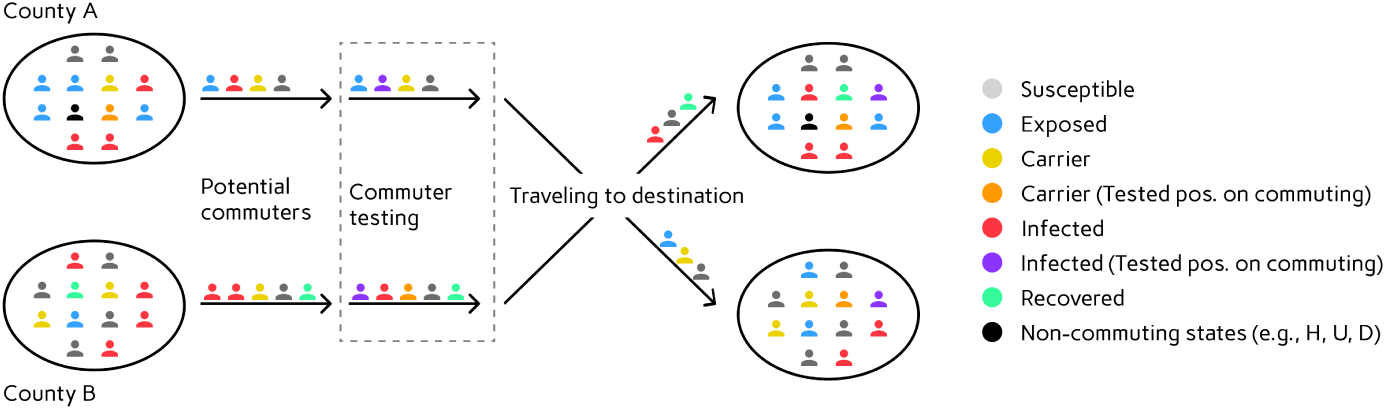
Implementation of commuter testing and traveling. Testing rates are applied according to the defined strategy and the states of the neighboring regions (red or green). Carriers or infectious who are tested positive become part of 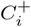 and 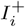 and are isolated accordingly. These individuals will not travel anymore before recovery.

For each age group *i* = 1, …, *n*, we denote the transmission risk by *ρ*_*i*_ and the proportion of carriers and infected people not isolated or quarantined is denoted by *ξ*_*C,i*_ and *ξ*_*I,i*_, respectively. The contact frequency matrix Φ = (*ϕ*_*i,j*_)_*i,j*=1,…,*n*_ represents the (mean) daily contacts of a person of age group *i* with people from age group *j*. For the remaining parameters, we use the variables 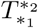 for the time spent in state ∗_1_ ∈ Ƶ_*i*_ before transition to state ∗_2_ ∈ Ƶ_*i*_ and 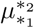 for the probability of a patient to go to state ∗_2_ from state ∗_1_.

Except for the transmission risk *ρ*_*i*_, the detection of carriers *ξ*_*C,i*_ and the isolation and quarantine of infected *ξ*_*I,i*_ defined in Table 1, we use the parameter ranges and age groups as gathered and described elaborately in [64, Table 2]. To account for the variant B.1.1.7 in Germany [3, Report of Apr. 7], we use a 1.4 times increased value for the transmission risk *ρ*_*i*_ [4]. The sigmoidal cosine curves in Table 1 are defined by the actual incidence of the zone. Thus, the minimum values are adopted for incidences below 12, where even carriers are quarantined and symptomatic are isolated fast and efficiently. From incidence 20 onward, carriers are generally no longer detected on a larger scale and the non-isolation of symptomatic increases to its highest value at incidence 150.

**Table 1.**
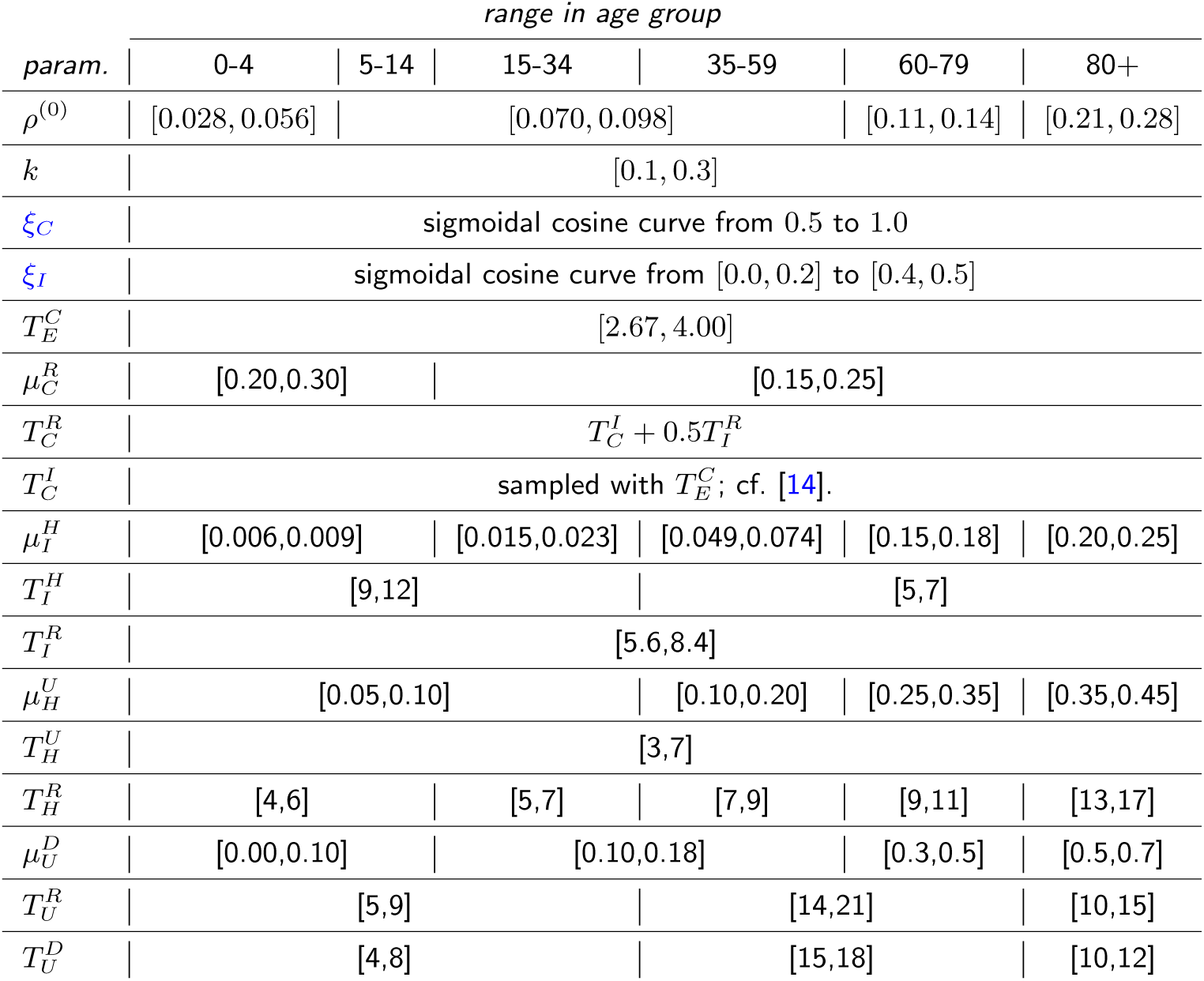
Parameter ranges used in our model. We omit the age index *i* for better readability. For derivation and more details, see [14].

**Table 2.** Description of parameters used in our model.

| Parameter | Description |
| --- | --- |
| $k$ | seasonality parameter for $s_k(t) := 1 + k \sin(\pi(\frac{t}{182.5} + \frac{1}{2}))$; cf. [14] |
| $\rho^{(0)}$ | baseline transmission risk |
| $\rho$ | transmission risk with seasonality effects: $\rho = s_k(t)\rho^{(0)}$ |
| $\xi_C$ | proportion of carrier individuals not isolated |
| $\xi_I$ | proportion of infected individuals not isolated |
| $T_E^C$ | period of latent non-infectious stage |
| $\mu_C^R$ | proportion of mild, asymptomatic cases |
| $T_C^R$ | period of asymptomatic stage before recovery |
| $T_C^I$ | period of latent infectious stage |
| $\mu_I^H$ | proportion of symptomatic cases needing hospitalization |
| $T_I^H$ | period of mild symptoms for individuals requiring hospitalization later on |
| $T_I^R$ | period of mild symptoms for individuals not requiring hospitalization later on |
| $\mu_H^U$ | proportion of hospitalized individuals getting ICU treatment |
| $T_H^U$ | period of hospitalization before ICU treatment (of critical cases) |
| $T_H^R$ | period of hospitalization before recovery (of non-critical cases) |
| $\mu_U^D$ | proportion of individuals in ICU care that die |
| $T_U^R$ | period of ICU treatment before recovery |
| $T_U^D$ | period of ICU treatment before death |

The range of contact patterns depends on the decreed non-pharmaceutical interventions. The baseline number of contacts *ϕ*_*B,i,j*_ is obtained from [91, 92] as described in [64, Sec. 3.2]. The resulting number of contacts according to the non-pharmaceutical intervention then writes

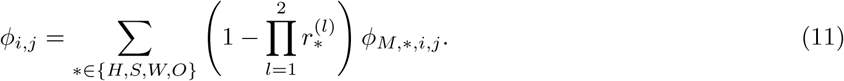

Here, ∗ ∈ {*H, S, W, O*} refers to the four locations of contact *home, school, work*, and *other*, and 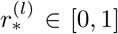 is the reduction factor in effective contacts as induced from political decisions. The superindex *l* is the intervention level. With *l* = 1 we describe interventions that yield direct contact reduction such as gathering bans. With *l* = 2 we include protective effects from, e.g., face masks and distancing; cf. [64] for more details. Precise values are provided in the following section. In contrast to [64], 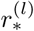 will not reduce the number of commuters from one region to another. The reduction in traveling of infectious and carriers will be based upon the isolation of symptomatic cases in the home region through 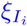 and the commuter testing rate as described in Fig. 2 and in the following section.

In order to account for the uncertainty, we consider an ensemble set of 500 Monte Carlo runs for each scenario such that our final results are based on 200 000 different runs.

### Mitigation and opening strategies

Our tensor space of strategies summarized in Table 3 provides 16 different strategies. These are defined by the dimensions *lockdown strictness* for red zones, *commuter testing rate* from red zones, and *delay of intervention implementation*. Political meetings of the German government with the federal state governments to discuss NPIs were often held in a four weekly rhythm, cf., [93, 94, 95, 96, 97, 98, 99, 100, 101, 102, 103, 104]. We thus assume that a set of restrictions always lasts for 30 days. It will only be lifted (red zones become green) if the incidence is below 12 at the end of the intervention.

**Table 3.** Tensor space of mitigation strategies. The 16 considered strategies are defined by choosing one item per column.

| Lockdown strictness | Commuter Testing | Delay of implementation |
| --- | --- | --- |
| 1.) **L+**: effective contact reduction:<br>70% ($I \leq 100$), 76% ($I > 100$) | 1.) **T0**: no testing | 1.) **D1W**: 1 week delay |
| 2.) **L**: effective contact reduction:<br>57% ($I \leq 100$), 64% ($I > 100$) | 2.) **T1**: 1 test per week | 2.) **D3W**: 3 weeks delay |
|  | 3.) **T2**: 2 tests per week |  |
|  | 4.) **T5**: 5 tests per week |  |

For all strategies, the handling of green zones is identical. We assume that all facilities can be opened and that first level political interventions are unnecessary, i.e., 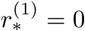 with ∗ ∈ {*H, S, W, O*} in eq. (11). However, second level interventions with face masks and distancing are in place, and we assume a reduced contact rate by 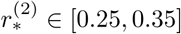 for ∗ ∈ {*S, W, O*} (schools, workplaces and other), and 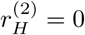 (homes), since face masks may not be worn in most private situations if the spread is under control.

For red zones, we consider two different sets of lockdowns. Both differ in their severeness for incidences below 100 and above 100. For the stricter set denoted by **L+** and incidences below 100, we implement first level interventions (*l* = 1) that achieve an average contact reduction by 50%. In addition (*l* = 2), we implement second level interventions with protective measures that increase the average effective contact reduction to 70% in total (*l* = {1, 2}). For incidences above 100, *l* = 1 leads to 58% and *l* = {1, 2} to 76% of effective contact reduction. For the less stricter set denoted by **L**, we have effective contact reductions (*l* = {1, 2}) of 57% and 64% with incidences below and above 100, respectively. For the Monte Carlo runs, we vary the contact reduction in a range of ±5%.

The particular contact reduction values for red and green zones and different locations (homes, schools, workplaces, and other) are based on [64] and on model calibration: red zones must substantially reduce their incidences in lockdown over multiple weeks and green zones are calibrated to maintain stable low incidence values within the local population (i.e. if there weren’t any commuters).

The second dimension of our strategy space is given by four different rates of commuter testing. With **T0**, there is no particular commuter testing at all, and with **T1, T2, T5** we refer to testing rates of once, twice, and five times a week. Based on a 5 days working week, **T5** will also be called the daily testing strategy. We assume massive deployment of antigen tests combined with a smaller number of PCR or RT-qPCR and pool tests [105, 106, 107] to control the commuter spread from highly infected regions. In [5], the sensitivity of antigen tests on the German market is estimated by 40-80%, and [108] identifies average chances of 72% and 58% to correctly detect an infection by antigen tests for symptomatic and asymptomatic cases, respectively. Given a combination of different kinds of tests, we use a generic daily detection ratio of 75% for carriers and infectious persons.

The last key property in our strategy space is the delay of implementation of new interventions once critical thresholds are exceeded. Since the introduction of NPIs always required political consensus, we decided to consider the effect of different delays from the moment an incidence threshold passed and a new intervention was decreed. We consider two well distinguished delays, a delay of one week denoted by **D1W** and a delay of three weeks denoted by **D3W** before new NPIs get active. This was done to study the effect of swift and slow reactions.

## Results

In this section, we present numerical results for the simulation of the 16 defined strategies for 25 randomized initial scenarios. In particular, we focus on the most effective strategy consisting of strict lockdowns, daily testing, and fast implementation of interventions (**L+, T5, D1W**).

In Fig. 3, we present four different scenarios of virus spread across Germany with 2 to 20 % of the regions classified as *red zones* (incidence 75-150 at start). These are generic scenarios of local outbreaks as they have been observed over the last year [3] in a smaller quantity. Each red zone represents a snapshot of the situation before the virus spreads into the surrounding regions. The initial situation is depicted on the top. In the center, we see the median outcome after 30 days and on the bottom the outcome after 60 days. Already after thirty days, we have a stable incidence of about 5 and only a small number of counties with incidences around 20 or 30. After 60 days, the initial heterogeneous situation is completely under control with stable incidences around 1 or 2 for all German counties.

**Figure 3.**
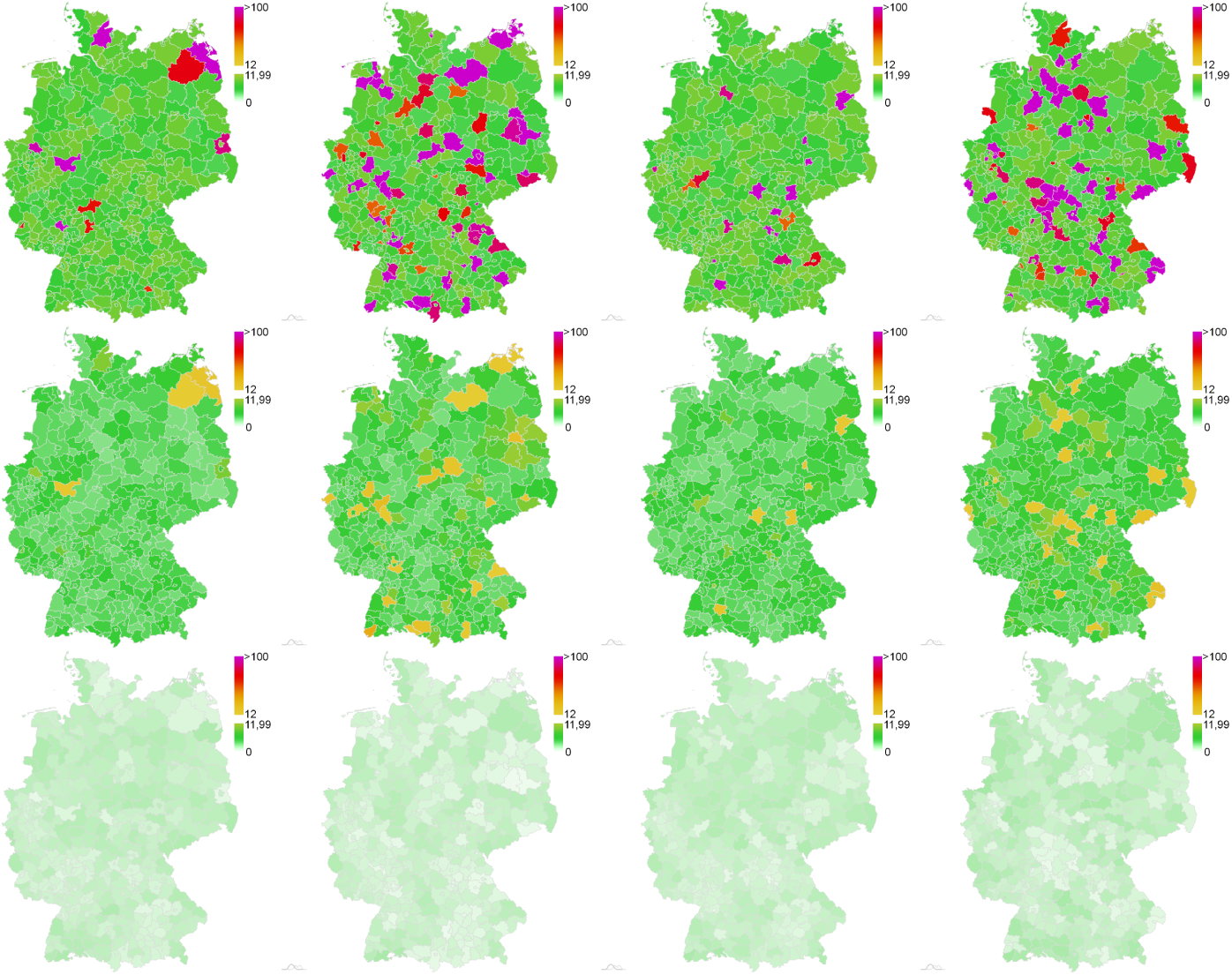
Simulated spread of SARS-CoV-2 for strategy **L+, T5, D1W** and four different initial scenarios (from left to right). Random initial incidence of 75-150 for 2-20% of the counties and incidence below 12 otherwise (top); state after 30 days (center) and after 60 days (bottom) of simulation. Median results from 500 Monte Carlo runs for each scenario.

In Fig. 4, we present the average incidence over the whole country for the 25 different initial red zone distributions. The median incidences (as well as the percentiles p25 and p75 of the Monte Carlo runs) for strategy **L+**,**T5**,**D1W** after 30 and 60 days are shown in orange and green, respectively. Analogous to the four scenarios considered in Fig. 3, the overall incidence drops considerably after 30 days already and is below 5 after 60 days in all 25 scenarios. As expected, a strong local lockdown **L+**, daily testing and a fast response time lead to green zones remaining green and red zones becoming green after only 1-2 months and thus avoiding a perpetuation of interventions.

**Figure 4.**
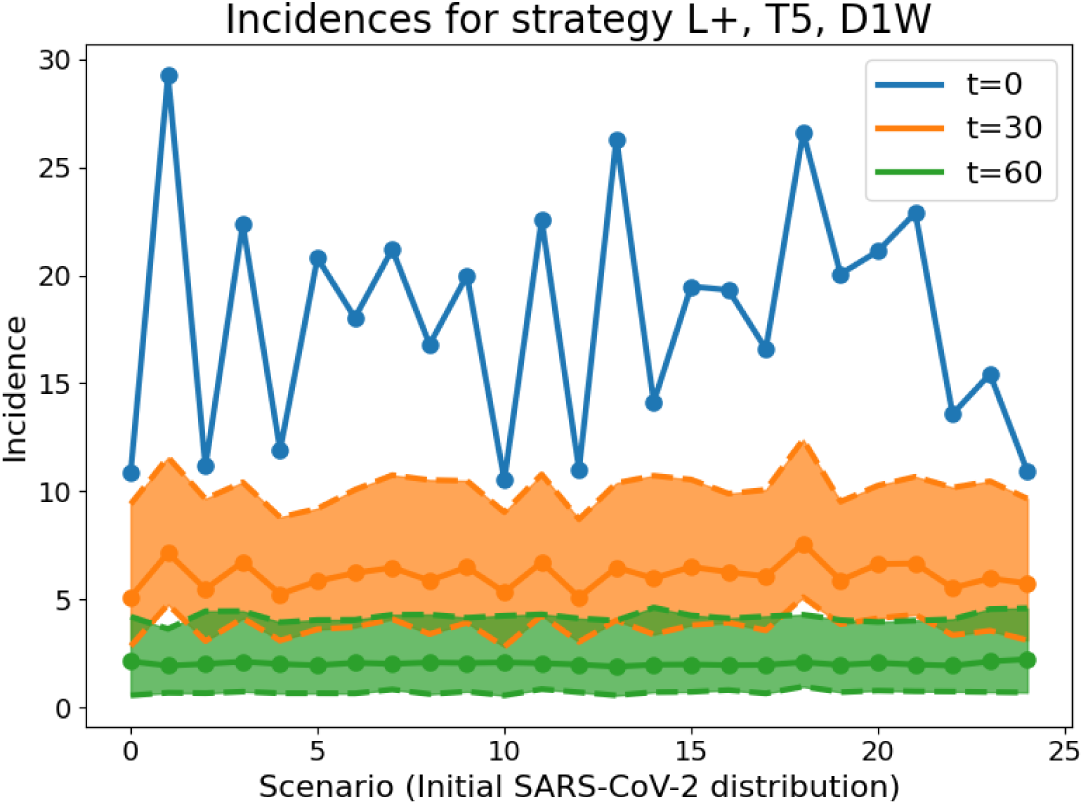
Country-wide SARS-CoV-2 infections per 100 000 and seven days (denoted: incidence) with the strongest mitigation strategy after 30 days (orange) and after 60 days (green) with the initial setting shown by the blue curve. The p25 and p75 percentiles for 500 Monte Carlo runs and 30 and 60 days of simulation, respectively, are shown by dashed lines in the same color.

In Fig. 5 and Fig. 6, we present the results for all strategies and all 25 initial scenarios. In Fig. 5, we see the number of counties in lockdown for all mitigation strategies (averaged over the 25 different scenarios) for 30 days of simulation time. Note that we use the same color for strategies that only differ in the strength of the lockdown **L+** (solid) vs **L** (dashed). In all cases, the **L+** strategy leads to less counties in lockdown over time. Hence, the most important factor for controlling the dynamics is the strictness of the lockdown in the red zones. The reduced lockdown strength **L** with an effective contact reduction of 57% (*I* ≤ 100) and 64% (*I >* 100) needs to be complemented by daily testing **T5** to control the propagation; see **L, T5, D1W** and **L, T5, D3W**. With **L** and lesser testing, the number of counties in lockdown doubles within just thirty days. Considering the slope of the curves, we see that the weaker the strictness of the interventions, the faster immediate action is required. With testing only once a week (**L, T1, D**∗**W**, ∗ ∈ 1, 3), almost a third of all counties will be in lockdown after only one month. In order to control the dynamics with testing rates under twice a week, very strict lockdowns **L+** with effective contact reductions of 70% (*I* ≤ 100) and 76% (*I >* 100) have to be implemented. These would have to be stricter than anything decreed for Germany ever before.

**Figure 5.**
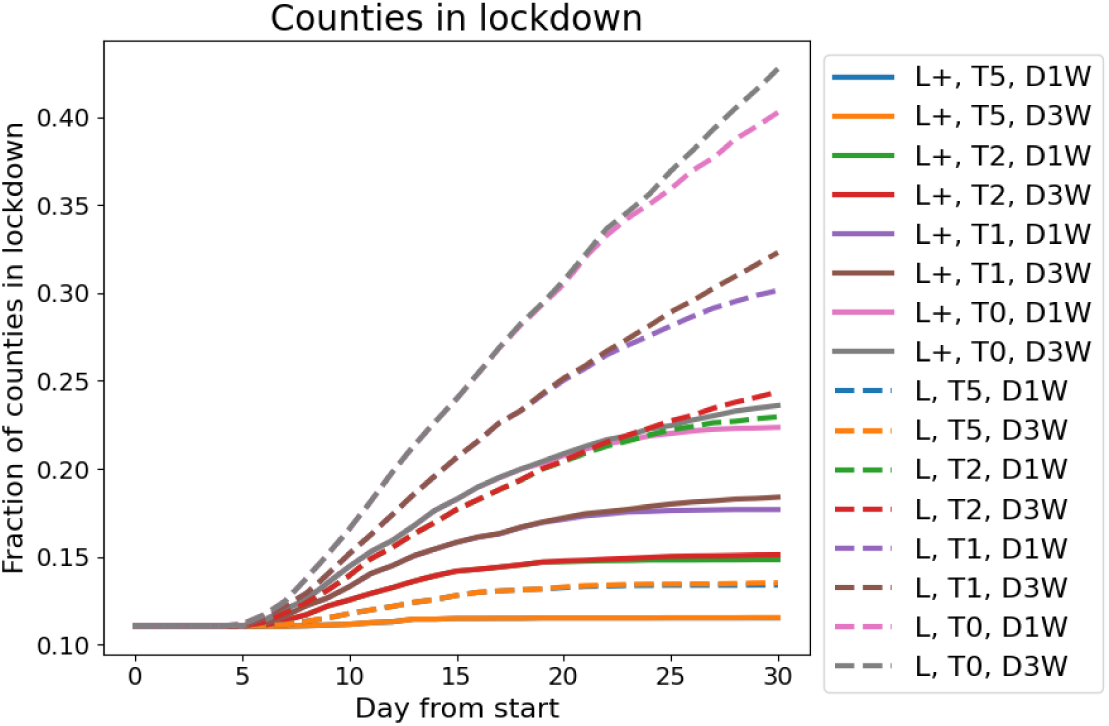
Counties in lockdown per day from start for the different mitigation strategies. Due to the strictness of the interventions in **L+, T5, D1W** and **L+, T5, D3W**, the delay of implementation is of minor importance and the curves overlap. For less strict interventions longer delays lead to more severe situations.

**Figure 6.**
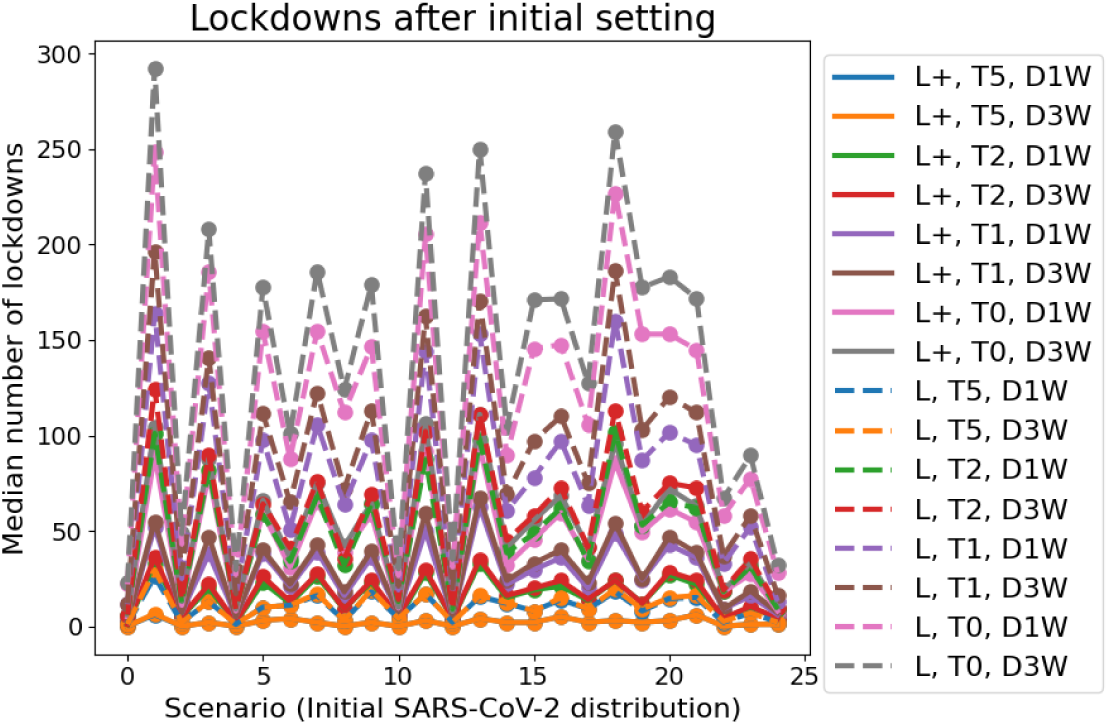
Median number of lockdowns per scenario (different initial distributions of SARS-CoV-2 spread) for the provided mitigation and opening strategies.

Fig. 6 depicts how many lockdowns have to be implemented for each scenario and each strategy over the whole country, i.e., how many counties turn red that were green in the beginning. Lesser restrictions like lockdown **L** and testing **T0, T1** or even **T2** lead to a substantial number of necessary lockdowns in neighboring regions. This number is 20-30 or even 100 times larger than that of strategy **L+, T5, D1W**. Stronger containment measures lead to more opening possibilities and hence to less economical damage than a perpetuation of less effective measures as also established in [82, 109]. The authors of [110] also advocated for aggressive political actions in the contagion containment phase to reduce the economic burden of the pandemic.

In Fig. 7, we depict the outcomes for one particular scenario of about 18 % red zones after 30 days of simulation with all 16 opening and mitigation strategies presented in Table Table 3. Again, strategies **L+, T5, D1W** and **L+, T5, D3W** lead to the most promising results. The images also quantify how reduced testing for the same lockdown strictness leads to slightly worse outcomes while reduced testing combined with reduced lockdown strictness quickly lead to out-of-control virus dynamics.

**Figure 7.**
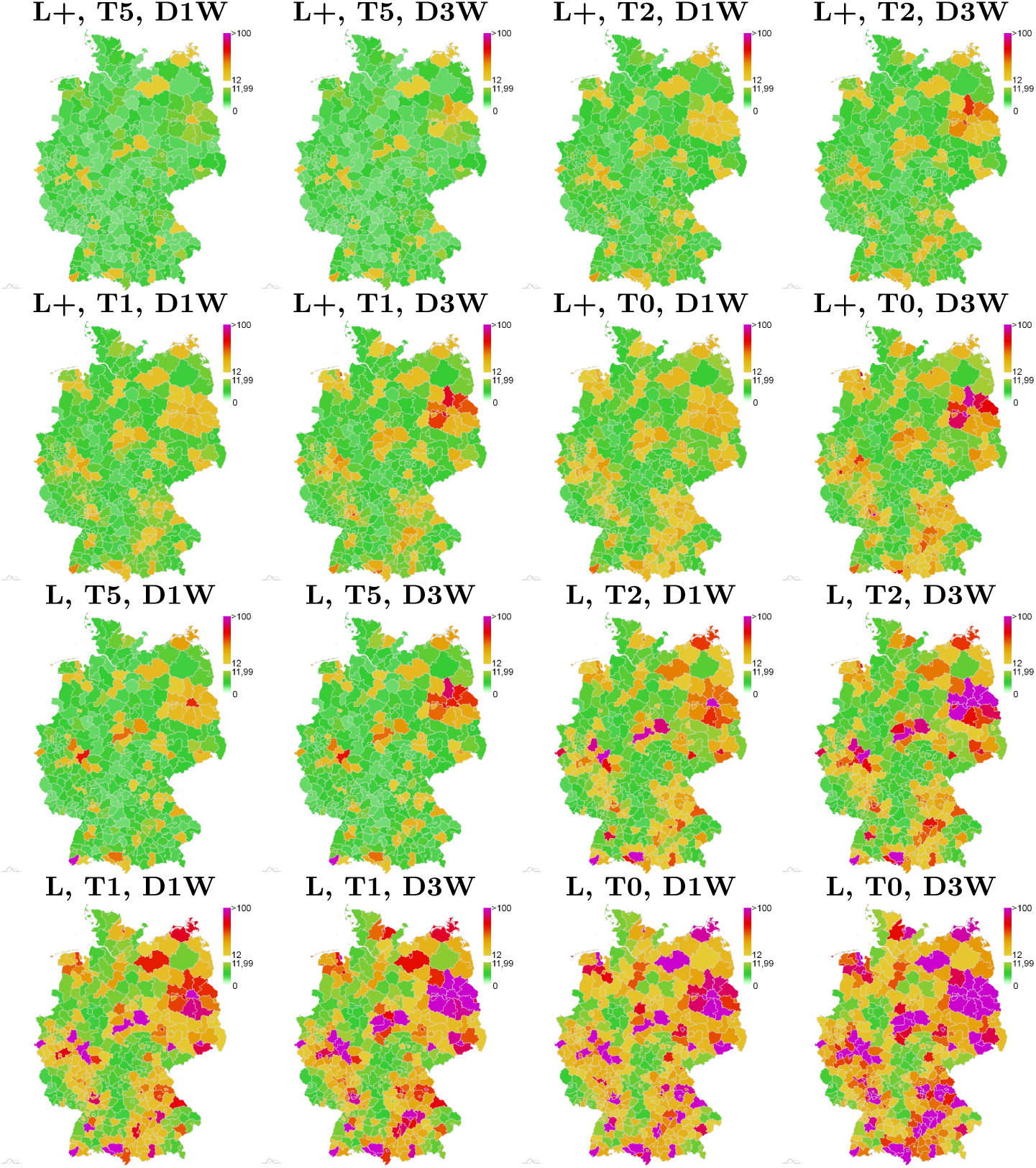
Simulated spread of SARS-CoV-2 cases for one initial scenario of about 18 % red zones and the 16 different strategies in Table 3. Each map represents the median result from 500 Monte Carlo runs after 30 days of simulation time. The incidence is computed per 100 000 and seven days. The maps are ordered according to the legend in Fig. 6 from **L+, T5, D1W** on the top left to **L, T0, D3W** on the bottom right. The initial distribution is the second to left scenario shown on the top in Fig. 3.

Finally, in Fig. 8, we present the outcome of the strongest and intermediate interventions, i.e. **L+, T5, D1W** and **L, T2, D3W**, for the case of a much severe initial situation with 42 % and 60 % counties as red zones. After 30 days with **L+, T5, D1W**, the situations improves considerably (center), while after thirty days with **L, T2, D3W**, we see a deterioration of infection dynamics (right).

**Figure 8.**
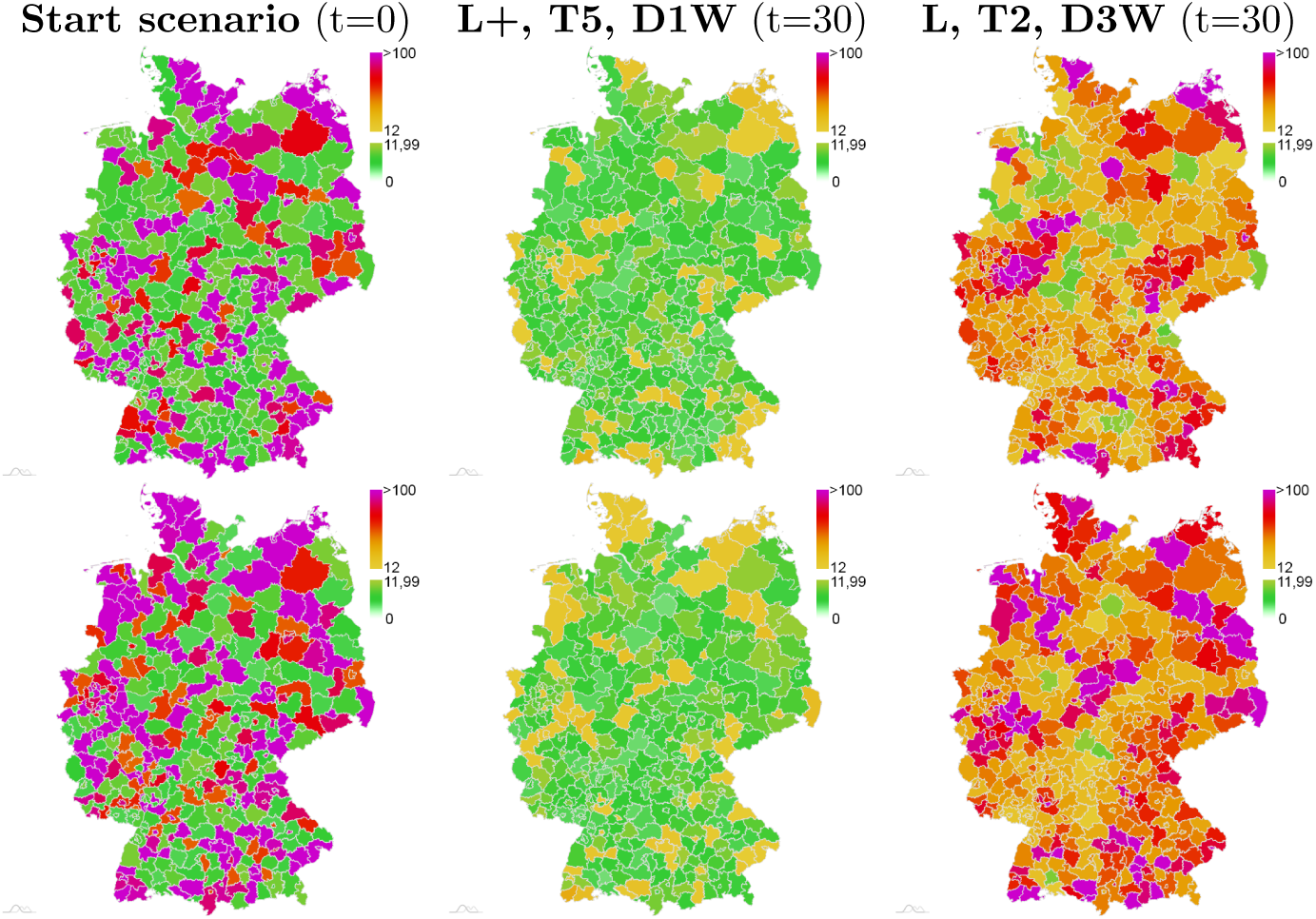
Two scenarios with 42 % (top) and 58 % (bottom) of counties classified red at initialization (left) and simulated spread of SARS-CoV-2 cases per 100 000 and seven days after thirty days of simulation with strongest strategy **L+, T5, D1W** (center) and intermediate strategy **L, T2, D3W** (right).

## Discussion

Within the first eighteen months of the pandemic, a variety of different approaches to controlling the spread of the SARS-CoV-2 pandemic has been proposed. Many authors tried to assess the effectiveness of different non-pharmaceutical interventions (NPIs). The authors of [109, 110, 111, 19] also partially focused on economic development and provided evidence for the effectiveness of lockdowns. The authors of [112, 113] derived confidence intervals for the effect of fine-grained NPIs such as school or workplace closures or gathering bans based on data for 149 and 41 countries, respectively. Although this assessment is an issue of high importance, this is not the focus of our paper. A key component in many successful interventions is a more or less differentiated and explicit zone distinction strategy based on regionally heterogeneous pandemic developments; see also Refs. [83, 84, 85]. Here, we investigated a basic version with only two different zones that are distinguished simply by incidence levels. While such zone distinction concepts are straightforward to implement in sparsely inhabited countries or countries with clear regional separation that facilitates isolation of high-incidence regions, the densely populated, highly mobile, highly connected situation in Germany, the EU, and similar regions presents challenges.

In particular, concepts of dealing with commuters between regions are needed. Work-related commuting between regions is often critical to maintaining a basic level of economic and social activity. Given the enormous cost of a complete ban on travel to neighboring cities, restricting mobility beyond a certain level seems unrealistic from a pragmatic point of view.

On the other hand, unrestricted travel between high- and low-incidence regions is problematic, as it effectively prevents regionally differentiated approaches due to the expected high amount of imported cases that will quickly revert any local progress towards lower incidence. Besides reducing unnecessary mobility between zones with different status to a minimum, testing commuters has been proposed as a tool to reduce import of cases into low-incidence zones. While this is intuitively appealing, its utility and practicality in a real-world scenario remains yet to be demonstrated. Recently, the authors of [111] found evidence in favor of domestic lockdowns to reduce the spread of the disease. On the other hand, the authors could not find any results in favor of border closures such that alternative strategies need to be found. Regular testing, however, may present a viable alternative that is more targeted and interrupts infection chains by isolating infected individuals.

In [114], 34 studies assessing the effect of fine-grained interventions were reviewed. While lockdowns were found to have an intermediate effect, testing was found to be less effective. However, when these studies were conducted, massive deployment of antigen tests was mostly not yet possible and effects of, e.g., daily or bi-daily testing in zones of high risk need to be studied. Also, interventions were often considered qualitatively and research on the interplay between the quantification of intervention strictness and testing is needed.

German health minister Karl Lauterbach recently declared that we still need strategies for ”hotspots”, i.e., regional outbreaks [115]. Our modeling approach aims at providing a data-based estimate of the effect that a combination of regionally differentiated restrictions and systematic testing of commuters with a given test sensitivity and frequency has on the overall incidence, on the frequency of necessary lockdowns, and on the containment of isolated local outbreaks. A limitation of the approach is that border regions are not considered, e.g., the impact of systematically higher incidences in a neighbouring country. However, since our simulation approach consists of randomly initialized generic scenarios, our conclusions also hold true for mobility across German borders.

Our results clearly indicate that a combined strategy of local lockdowns and systematic testing has the potential to contain isolated local outbreaks in a general low incidence setting. In particular, rather strict local measures in combination with frequent testing of commuters proves highly effective in preventing the spread of localized infection hot spots and reducing the incidence in these regions. This can be seen both in the number of counties that need to impose measures and in the median incidence in the simulations which can be brought to surprisingly low levels with the most effective strategies.

The results further clearly indicate a hierarchy of effectiveness that depends on the strength of the locally imposed measures, the duration of the delay with which they are imposed after a critical incidence threshold has been reached, and the frequency at which commuters are tested which ultimately determines the fraction of imported cases that are found.

Interestingly, although the strength of the locally imposed measures dominates the further development of a local outbreak as expected, our simulations indicate that with an effective daily testing regimen and a swift reaction to local outbreaks, even a less severe reaction may be effective in containing isolated outbreaks. With less strict contact reduction policies in the lockdown scenario, our model predicts especially testing frequency to be a highly relevant factor in outbreak mitigation. It should be noted, however, that our scenarios are low-incidence scenarios with a rather small number of local infection hot spots. With a high overall incidence as demonstrated in Fig. 8, strict measures are even more essential in first instance. With very strict policy interventions, reaction delay and testing of commuters appear to have less of an impact. However, testing only once a week or not at all can quickly lead to a degeneration of infection dynamics.

## Conclusions

Using a regionally resolved model based on mobility data to describe pandemic spread on a subnational level, we systematically investigated the feasibility of a localized strategy for outbreak containment based on a simple distinction of low- and high-incidence regions. Such a strategy may be especially useful in a scenario where generalized, nation-wide measures have brought incidence to a rather low overall level and a restrictive overall policy is no longer necessary. In that sense, it can be seen as a perspective for careful resumption of economic and social activity. This is especially important as there is a clear general pattern of re-emergence of the pandemic after successful containment and lifting of restrictive policy; e.g., the incidence in Germany remained at very low levels after the first outbreak in March 2020 until September when several local outbreaks emerged and, after lack of a substantial intervention, developed into a nationwide second surge of infections. Similar developments could be observed after Summer 2021.

Our results indicate that local containment of outbreaks and maintenance of low overall incidence is possible even in densely populated and highly connected areas. While we demonstrate this on data from Germany, similar patterns of mobility likely exist in many countries and our results are, hence, generalizable to a certain extent.

While it is obvious that a substantial reduction of transmission and, hence, contacts in the population is necessary to mitigate a local high-incidence situation, our results suggest that reduced mobility along with frequent testing of regular commuters can successfully prevent generalized spreading of the infection in larger areas even with less strict contact reduction policies, especially when reaction to an outbreak is swift. This gives rise to a promising perspective after hard and economically damaging policy interventions: Maintaining the situation at stable levels may require moderate, localized interventions that affect only a small fraction of the population, offering a viable alternative to switching back and forth between premature lifting of restrictions and restrictive untargeted measures.

## Data Availability

The parameters to setup the simulations are provided in the current article and [64]. The simulation output for all different strategies, scenarios and Monte Carlo runs presented here make up more than 100 GB in over 1.9 million items and cannot be shared easily. Particular results can be shared upon request. Please contact. Intra- and intercounty contact and raw, non-normalized mobility data for initialization are provided in text files. The underlying software framework MEmilio has been published and is available as open source on https://github.com/DLR-SC/memilio.

## Declarations

## Abbreviations

Covid-19: Coronavirus disease 2019
SARS-CoV-2: severe acute respiratory syndrome coronavirus type 2.

## Ethics approval and consent to participate

Not applicable.

## Consent for publication

Not applicable.

## Acknowledgements

Not applicable.

## Authors’ contributions

M.M.-H., S.B., M.J.K. and M.K. developed the mitigation strategies, M.J.K., D.A., M.K., J.K., W.K. and M.S. developed and implemented the graph SIR-type model with commuter testing, M.J.K., K.R. and W.K. developed the data analysis tools, J.G. and L.S. developed the visualization tools, M.M.-H. and A.B. supervised the study.

## Additional Files

Contact and mobility raw data — raw mobility contact.zip

This files includes the baseline intra-county contact patterns and the raw, non-normalized mobility data, not yet differentiated for age-groups, used for initialization of inter-county mobility.

## Footnotes

[1] The small deviation from the incidence 12 in [82] takes into account reporting delays.

